# A Th17 signature correlates with response to dual CTLA-4 and PD-1 blockade in metastatic renal cell carcinoma

**DOI:** 10.1101/2025.09.29.25335360

**Authors:** Silvia Guglietta, Anne Bärenwaldt, Anthony Sonrel, Stephany Orjuela, Gianni Monaco, Thomas Cahill, Petra Herzig, Alfred Zippelius, Lixia Li, Lukas Bubendorf, Robert Ivanek, Gary Hardiman, Vanessa Peterson, Mark D. Robinson, Frank Stenner, Heinz Läubli, Carsten Krieg

## Abstract

The advent of combination immune checkpoint inhibitors (cICI), specifically ipilimumab and nivolumab, has transformed the treatment landscape for metastatic renal cell carcinoma (mRCC), offering durable remissions and improved outcomes for intermediate- and poor-risk patients. However, intrinsic resistance remains a significant challenge, with 40-60% of patients failing to achieve meaningful responses. Here, we conducted a comprehensive, multi-omic analysis of systemic and tumor-associated immune responses in mRCC patients enrolled in a clinical trial (CA209-980, SAKK07/17, NCT03297593) testing a novel response-adapted cICI regimen. Our study aimed to identify immune correlates of response to cICI therapy.

High-dimensional mass cytometry and single-cell proteomic and bulk RNA sequencing of the tumor revealed an enrichment of Th17 CD4+ T cells in responders. These cells exhibited upregulated IL-21-driven pathways, IL-17 signaling, and inflammasome-associated processes, highlighting their central role in therapeutic efficacy.

Our integrative analysis underscores the importance of Th17 cells in mediating effective responses to cICI and provides a framework for developing predictive biomarkers and therapeutic strategies to overcome resistance. These findings highlight the translational potential of targeting Th17-centric pathways to enhance immunotherapy outcomes in mRCC and potentially other cancers. This work represents a significant step toward advancing personalized oncology through integrated immune profiling.

## Introduction

The treatment of metastatic renal cell carcinoma (mRCC) has significantly improved during the last years following the introduction of anti-angiogenic therapy and immune checkpoint inhibition (ICI)^1-4^. The combined ICI (cICI) with anti-PD-1 (nivolumab) and anti-CTLA-4 (ipilimumab) antibodies has shown impressive results, particularly in patients with intermediate and poor risk suffering from metastatic clear cell carcinoma, where ICI has shown a high rate of complete response and durable remissions^5,6^ compared to monotherapy or anti-angiogenic agents such as the standard of care Sunitinib^2,6^. In addition, studies have also shown efficacy of cICI in patients with sarcomatoid features, who have notoriously a very poor prognosis^7^. Despite these encouraging data and the improvement in quality of life, cICI therapies result in immune related adverse events that affect the endocrine system, are often irreversible and may result in therapy discontinuation with consequent loss of therapeutic benefit. Furthermore, 40–60% of patients still have intrinsic resistance to ICIs across multiple clinical trials and often require further therapy^2,6^. Therefore, there is a significant unmet need in the field for improved response stratification and combination approaches such that we can extend the benefits of cICI to larger groups of patients affected by mRCC. Tumor responses to therapy with substantial tumor shrinkage (partial response, PR, defined as >30% tumor reduction by RECIST assessment or complete response, CR) are associated with a better prognosis in patients with advanced RCC^1-4^. Even more important, depth of response has been associated with long-term survival^8^.

Response biomarkers and cellular phenotypes associated with response are objects of active investigations in the field^9,10^. Mutations in *PBRM1* in cancer cells were initially associated with worse outcomes in RCC patients treated with ICI^5^. However, more recent and more extensive investigations have showed no correlation between tumor molecular features and response^11^. More recently, there has been a growing interest in understanding the role of immune cells in response to ICI in mRCC. While these studies have provided invaluable information on markers, cell types and immune pathways during ICI treatment in mRCC, they also suffer from several limitations. Namely, most of the published data are from patients treated with single agent ICI (anti-PD1). Furthermore, the focus on PD-L1 expression in the tumor microenvironment (TME) and on the tumor mutational burden (TMB), which have proven successful in predicting response to ICI treatment in many cancers, yielded disappointing results in the stratification of mRCC patients^12-14^. Similarly, in contrast to many other cancer types, the notable infiltration of CD8+ T lymphocytes in RCC does not necessarily translate into improved treatment responses or prognosis in mRCC^15^. Other studies using more unbiased methods showed that high T cell/low myeloid infiltration and high B cell abundance are enriched in responders to anti-PD-L1 or PD-1^16,17^. However, cross-validation of these features was inconsistent possibly because these studies solely relied on tumor samples^5,18^.

To address the significant unmet need for improved stratification and therapeutic strategies in mRCC by defining immune response signatures, we comprehensively examined the immune responses to cICI in patients with poor/intermediate risk mRCC enrolled in a clinical trial (CA209-980, SAKK07/17, NCT03297593) testing a novel regimen with nivolumab (anti-PD1) and a response-adapted ipilimumab (anti-CTLA-4) dosing schedule designed to reduce adverse events while preserving efficacy. While the clinical results are part of a separate study, our approach focused on identifying systemic and tumor-associated immune correlates of responses. Through high-dimensional mass cytometry, we evaluated T cell and non-T-cell populations in peripheral blood. Transcriptomic changes in bulk tumors and metastases provided a detailed map of immune pathways. A systems immunology approach integrated these data, revealing a strong Th17-centric signature linked to therapeutic responses. These findings highlight the potential of Th17 cells as both biomarkers and mediators of therapeutic success, offering a foundation for enhancing precision oncology in mRCC.

## Materials and Methods

### Patients and treatment intervention

The Checkmate 209-980 (SAKK 07/17) trial is a phase II, single-arm clinical trial testing an alternative schedule of ipilimumab and nivolumab immunotherapy for advanced renal cell carcinoma. Patients with histologically or cytologically confirmed, locally advanced, and/or metastatic clear cell RCC not amenable to surgery or definitive radiotherapy and requiring systemic immunotherapy were enrolled. Patients started treatment with nivolumab (240 mg every two weeks during the first 20 weeks, 480 mg every four weeks thereafter). After two weeks, ipilimumab (1mg/kg every six weeks) was introduced. As soon as a radiographic complete response (CR) or partial response (PR) was observed, ipilimumab was stopped, and treatment with nivolumab continued. The main endpoint of this phase II trial was the progression-free survival (PFS) after 12 months.

### CyTOF (cytometry by time of flight) on peripheral blood cells

Individual samples of up to 3×10^6^ cells were barcoded with a combination of platinum-conjugated or indium-conjugated anti-human β2m and anti-CD298 antibodies in cell staining buffer (CSB; PBS, 5mM EDTA, 2% BSA) for 20min at 4°C as described previously^19^. A unique to the sample seven-choose-three barcoding matrix was used to allow for sample surface staining in one tube and doublet removal. Cells were washed twice in CSB and combined in one tube for surface stains. In preparation for the viability stain, cisplatin (Sigma) was resuspended in DMSO, pre-conditioned for 48h at 37°C, aliquoted, and stored at -20°C. Cells were washed in PBS, and the viability staining was done with 200μM Cisplatin for two min in PBS at room temperature (RT). All surface staining was performed in CSB for 30 min at RT, and cells were fixed in 2%PFA for 10 min at 4°C. Before the acquisition, cells were resuspended in iridium intercalation solution (2%PFA, 0.02% saponin (Sigma), and 0.5mM iridium in PBS) overnight at 4°C. For data, acquisition samples were washed once in CSB and twice in ddH2O. Cells were filtered through a cell strainer, resuspended in ddH20 containing 1xEQ beads at a density of 1×10^6^/ml, and acquired on a Helios mass cytometer (Standard BioTools).

### CyTOF Data Analysis

CyTOF data was loaded and pre-processed in R with the CATALYST R package^20^. Cells were clustered based on 36 protein-type markers for T cells and 35 for non-T-cells using FlowSOM^21^, and clusters were manually annotated using the protein expression of the same markers (**Supp. Tables 1 and 2**). Differential abundance between cell clusters was detected with diffcyt^22^ accounting for patient and sample variability. To identify differences in single-cell protein expression or cellular abundance in responder patients, we used mass cytometry and designed a 37-marker panel targeted at T cell and one 35-marker panel targeted at non-T-cells. Based on previous work by Sallusto et al., we used a set of chemokine surface markers (CCR4, CCR6, CCR7, CXCR3, CD127, CCR7, CD62L, CD28, CD45RO, CD57, CD95) to define T cell subsets^23^. All samples were acquired at the same time in one batch.

### Bulk RNA sequencing of primary tumors

Biopsies were homogenized in Tri-Reagent (Sigma) and stainless-steel beads (Qiagen) using the TissueLyser 2 (Qiagen). RNA extraction was performed using the Direct-Zol RNA Kit (Zymo Research) according to the manufacturer’s instructions. RNA were quantified by Fluorometry using the QuantiFluor RNA System (Cat# E3310, Promega, Madison, WI, USA) and were quality-checked on the TapeStation instrument (Agilent Technologies, Santa Clara, CA, USA) using the High Sensitivity RNA ScreenTape (Agilent, Cat# 5067-5579), which showed an average RIN of 7.6. Library preparation was performed starting from 50ng total RNA using the TruSeq Stranded mRNA Library Kit (Cat# 20020595, Illumina, San Diego, CA, USA) and the TruSeq RNA UD Indexes (Cat# 20022371, Illumina, San Diego, CA, USA). 15 cycles of PCR were performed. Libraries were quality-checked on the Fragment Analyzer (Advanced Analytical, Ames, IA, USA) using the Standard Sensitivity NGS Fragment Analysis Kit (Cat# DNF-473, Advanced Analytical). Average concentration was 66±14 nmol/L and average library size was 177±2 base pairs). Libraries were pooled to equal molarity. The pool was quantified by Fluorometry using the QuantiFluor ONE dsDNA System (Cat# E4871, Promega, Madison, WI, USA). Libraries were sequenced Paired-End 51 bases (in addition: 8 bases for index 1 and 8 bases for index 2) using the NovaSeq 6000 instrument (Illumina) and the S1 Flow-Cell loaded at a final concentration in Flow-Lane of 380pM and including 1% PhiX. Primary data analysis was performed with the Illumina RTA version 3.4.4. The enrichment score was calculated with the Singscore method (v. 1.22), which uses rank-based statistics, on log2-transformed TMM values. Heatmap was generated with the pheatmap package (v. 1.0.12). Boxplots were generated with ggplot2 (v. 3.5.0) and a t-test with associated visualization was performed with ggpubr (v. 0.6.0).

### Pathway Visualization and Analysis Using Cytoscape

Gene Set Enrichment Analysis (GSEA) (v4.3.3) was conducted on bulk RNA sequencing data from responder and non-responder groups to identify enriched pathways. Preprocessed gene expression data were ranked, and GSEA results were assessed using Normalized Enrichment Scores (NES) and p value (p ≤ 0.05). GSEA results were visualized in Cytoscape (v3.10.2) using the EnrichmentMap plugin (v3.4.0), displaying enriched gene sets as nodes and their overlap as edges. Genes differentially expressed in CD4+, CD4CM, and CD4TH17 T cell subsets and involved in immune processes were mapped onto the EnrichmentMap to link gene-level changes with enriched pathways and biological processes in the blood and tissue.

## Results

### Single-cell proteomic analysis shows an association of Th17 cells with antitumor immunity following cICI

This study was initially designed to include 37 patients; however, due to encouraging early clinical activity, enrollment was expanded to a second cohort, resulting in a total of 74 patients treated within the context of an interventional phase 2 trial (CA209-980/NCT03297593/SAKK 07/17). The clinical trial’s primary endpoint was the assessment of toxicity and safety of a novel combination of anti-CTLA-4 and anti-PD-1 in patients with intermediate- and poor-risk mRCC. While detailed toxicity data and treatment outcomes will be reported in a separate clinical manuscript, the current report focuses specifically on translational analyses designed to identify immune correlates of response.

These exploratory biomarker studies were conducted on the first cohort of patients, from whom serial peripheral PBMCs as well as matched pre- and on-treatment tumor tissue (primary and/or metastatic) were available. Paired blood and tumor samples were successfully collected from 24 patients and form the basis of the translational analyses presented here. Among these patients, 16 experienced either a partial or complete response (PR, CR), while 8 had stable or progressive disease (SD, PD) (**Supp. Table 3**). In addition, PBMCs from two healthy donors were included for selected comparative analyses. These specimens enabled an in-depth evaluation of immune mechanisms associated with response or resistance to cICI therapy in mRCC.

We are aware that possible gene signatures could support the response to immunotherapy in ccRCC, particularly in the metastatic setting. Prior studies have suggested that loss-of-function mutations in *PBRM1* are associated with improved responses to immune checkpoint inhibitors^24^, while alterations in *BAP1, PBRM1*, and metabolic pathways have been linked to decreased survival across RCC subtypes^25^. However, more recent data on metastatic RCC have shown conflicting results regarding *VHL, PBRM1*, and *SETD2* mutations, with *SETD2* alterations potentially predicting poorer outcomes and concomitant *VHL* and *PBRM1* mutations possibly enhancing ICI/TKI efficacy^26^. Despite these insights, our own analysis did not find a correlation between *VHL, BAP1, PBRM1*, or *SETD2* and immunotherapy response in our patient cohort, highlighting the need for further large-scale validation of therapeutic predictors (**Supp. Fig. 3**).

Adaptive immune cells are the major target in immunotherapy, but the contribution of non-T cells to therapeutic success is increasingly recognized. Unfortunately, studying a significant amount of innate and adaptive immune cells at the same time carries the challenge of an about five-fold higher abundance of adaptive immune cells compared to innate immune cells in PBMC. To study both cell populations equally, we used pan-anti-TCR-beta antibodies to magnetically separate blood T cells from other less abundant PBMC (**Supp. Fig. 1**). To minimize the batch effect, we used barcoding or hashing and pooled all samples before further processing (**Supp. Fig. 1**). To discover differences in cellular abundance and protein expression, we first used mass cytometry and designed two antibody panels. First, we analyzed the T cell fraction in the peripheral blood based on protein expression alone (**Fig. 1A, Supp. Table 2**). After normalization, we examined the effect of therapy on differential protein marker expression. No differences in overall protein marker expression were found between treatment groups before or after therapy (**Supp. Fig. 2A**). Using the normalized protein expression matrix of each cell fraction, we performed Louvain clustering to identify cell clusters driven by protein expression. We identified and manually annotated nine distinct clusters based on their relative protein expression. Clusters included B cells, γδ T cells, CD4 T cell, and CD8 T cell subpopulations. Following cell type annotation, we observed an increase in the abundance of naïve CD4 T cells (CD3^+^, CD4^+^, CD45RA^+^, CD45RO^-^, CCR7^+^), Th17 cells (CD3^+^, CD4^+^, CCR4^+^, CCR6^+^), CD8 effector memory (EM) T cells (CD3^+^, CD8^+^, CD45RO^+^, CCR7^-^) and γδ T cells (CD3^+^, Tcr-γδ^+^) in response to the cICI treatment (**Fig. 1A**). Of note, the frequencies of naïve CD4 T cells and γδ T cells were also significantly elevated at baseline. We next used the second panel (**Supp. Table 3**) to investigate the non-T-cell fraction (**Figure 1B**). Similar to the T cell fraction, we found no difference in the protein abundance and identified nine cell populations (**Supp. Fig. 2B**). Interestingly, several clusters in the non-T-cell fraction showed differential abundance. Specifically, as shown in **Fig. 1B**, we observed higher percentages of cDC1, cDC2, and preDC sub-populations after cICI treatment in responders. We did not find significant differences in cell abundance between responders and non-responders before therapy.

**Figure 1.**
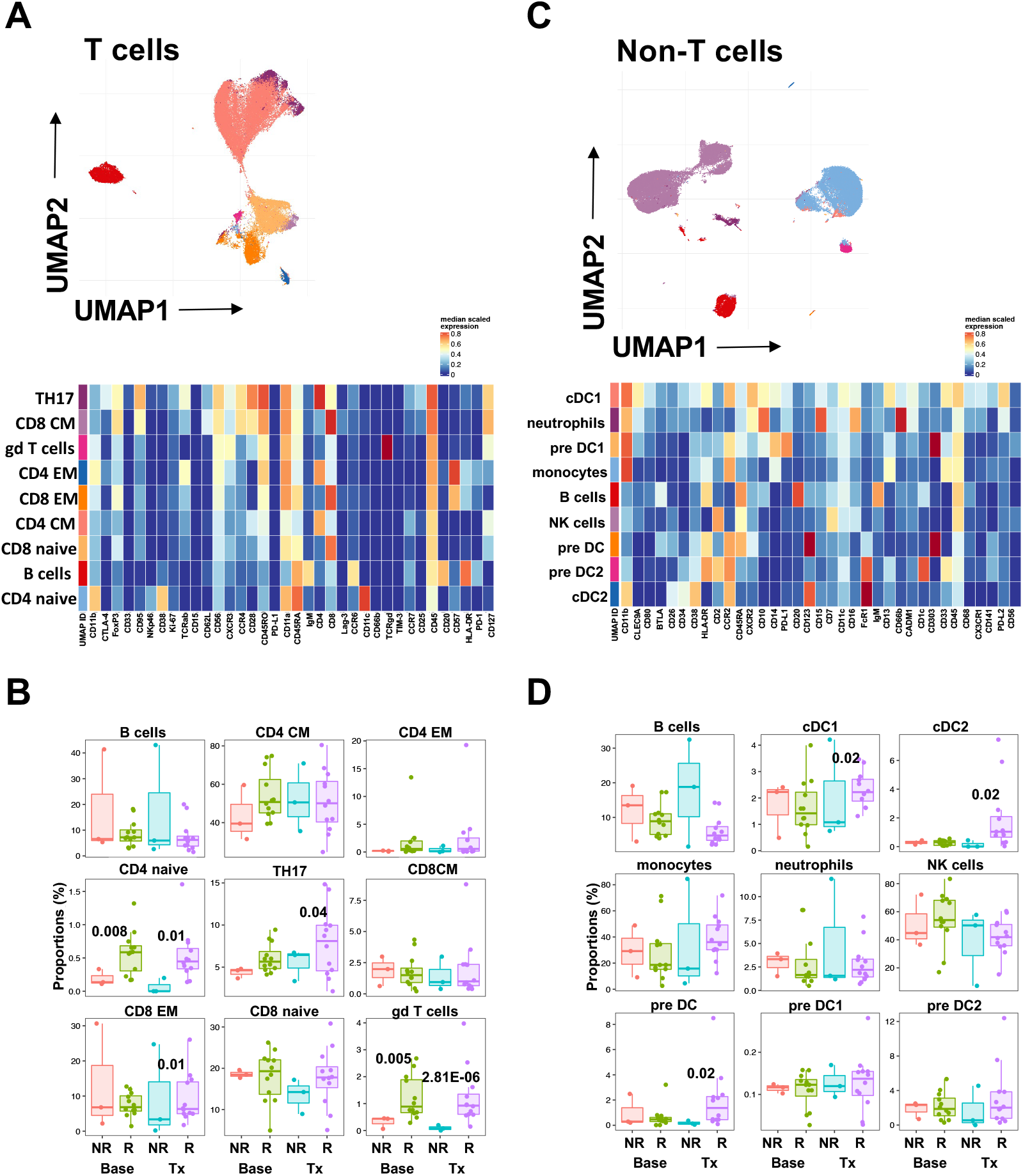
Characterization of peripheral blood PBMCs by CyTOF. **A**, Top left, UMAP visualization of FlowSOM-generated T cell clusters and heatmap of the expression values of the indicated markers within the cellular clusters. **B**, Differential abundance of T cell subsets from responder patients before and on therapy. **C**, UMAP visualization of FlowSOM-generated non-T-cell clusters and heatmap representing the expression of the indicated markers within the cellular clusters. **D**, Differential abundance of non-T-cell subsets from responder patients before and on therapy. Control (Ctr, red), non-responder (NR, green), and responder (R, blue). Heatmap display the normalized (a mean of 0 and a s.d. of 1) median expression for markers. Displayed are adjusted p-values between responder and non-responder were significant. Box plots represent the IQR, with the horizontal line indicating the median. Whiskers extend to the farthest data point within a maximum of 1.5× IQR.

Altogether, our discovery approach using CyTOF revealed expansion of Th17, CD8EM cells and DC subsets in the peripheral blood of responder patients following cICI treatment.

### Analysis of transcriptomic profiles in tumor biopsies shows changes in Th17 cells in responders that mirror the findings in the peripheral blood

We performed bulk RNA sequencing on selected baseline tumor specimens. 31 biopsies were sampled from different tissue sites in responders and non-responders. These included the primary kidney tumor (sometimes also excisions) and biopsies of metastases from liver, bone, lung, soft tissue, pancreas, and lymph nodes. Using our previously identified gene expression profiles from T and non-T-cell single-cell analysis in the peripheral blood, we probed the tumor transcriptome to test for the presence of similar signatures. As shown by the z-score marker expression in the heatmap (**Fig. 2A)** and the SingScores (**Fig. 2B**), we found an enrichment in Th17 cells, CD4EM, and CD4E gene signatures in responder vs non-responders. Importantly, the results were not dependent on the site of the sampled biopsy. Next, we compared our newly identified transcriptomic signature to a previously published interferon-responsive gene signature after immunotherapy in over 300 samples and over 22 cancer types^27^. While we observed some overlap, the Th17-centric tumor signature is a unique finding from our study. Overall, this analysis on tumor biopsies confirmed that our CD4-centric and Th17 gene signature identified in the peripheral blood of mRCC patients is also found at the tumor site and correlates with response to cICI.

**Figure 2.**
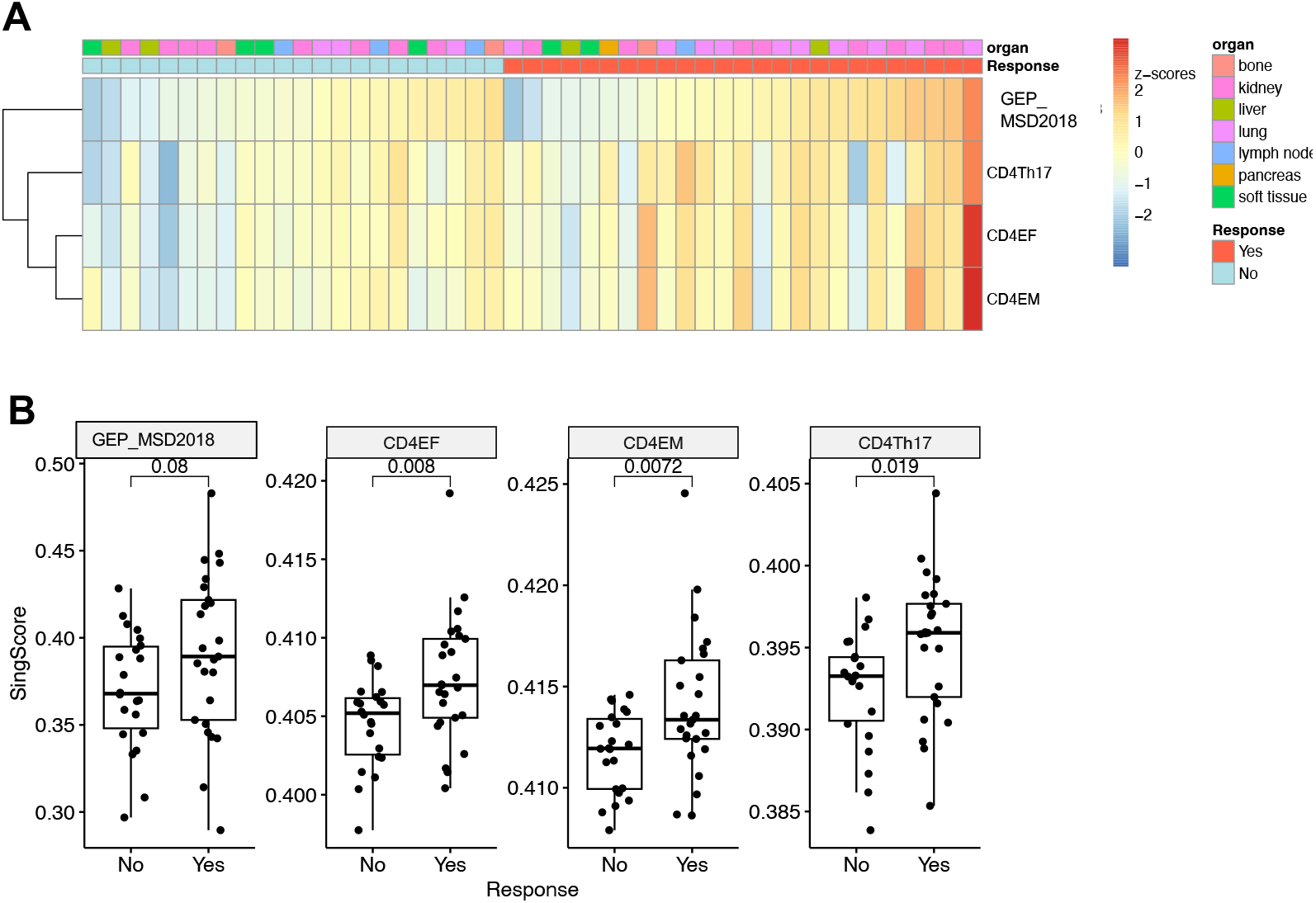
Enrichment scores of T cell signatures related to anti-tumor response. **(A)** Heatmap shows the z-scores of the enrichment scores from transcriptomics data of different tissue biopsies of three T cell types (CD4 Th17, CD4EF, CD4EM) as well as a signature previously found to be overexpressed in responders (GEP_MSD2018). **(B)** Box plots showing the original enrichment score values and significance obtained by t-test analysis.

### Identification of specific molecular pathways in the TME

Despite the low tumor mutational burden (TMB) in RCC - approximately tenfold lower than melanoma and comparable to “cold” tumors - RCC displays unique immunological features. These include high immune infiltration scores and increased infiltration by cytotoxic CD8+ T cells, which are typically associated with favorable responses to PD-1 blockade in other cancers such as NSCLC, melanoma, and microsatellite instability-high (MSI-H) colorectal cancer^25,28^. However, unlike other cancer types, this pronounced CD8+ T cell infiltration in RCC does not necessarily correlate with improved treatment response or prognosis in mRCC^29^.

Therefore, in our study we mainly focused on CD4+ T cells as potential mediators of antitumor responses. CD4+ T cells are central to orchestrating immune responses, influencing other immune cell populations through cytokine production and pathway activation. Recent evidence suggests that subsets of CD4+ T cells, such as Th17 cells, may play pivotal roles in shaping both systemic immune responses and the TME in response to ICI. We, therefore, sought to investigate systemic and local therapeutic responses by analyzing CD4+ T cells in peripheral blood and tumor samples to uncover shared and unique pathways driving treatment efficacy.

To elucidate the molecular underpinnings of immune responses in the TME contexts bulk transcriptomic data was generated from tumor samples. Data integration and pathway analysis were conducted using the Advaita software platform, allowing for comprehensive examination of hallmark pathways and key genes driving immune responses across both compartments.

The analysis incorporated pathway and annotation data from multiple databases, including KEGG (Release 110.0+/05-01, May 2024) for functional pathway insights, Gene Ontology (2023-Sep-11) for molecular function and biological process classifications, miRNA-target interactions from miRBase (Version 22.1, October 2018) and TARGETSCAN (Version 8.0), regulatory networks from BioGRID (Version 4.4.233, April 2024), and chemical-disease interactions from the Comparative Toxicogenomics Database (October 2023). Additionally, disease associations were sourced from KEGG to contextualize identified pathways. This integrative strategy enabled robust exploration of biological and regulatory interactions relevant to immune modulation.

In total, we analyzed 1,876 genes from peripheral blood and 22,704 genes from tumor samples. Differential expression analysis revealed 466 differentially expressed genes (DEGs) in blood (top 5%) at a threshold of ≥0.6-fold change (p < 0.05) and 1,247 DEGs in tumor samples (top 20%) at a threshold of ≥1.2-fold change (p < 0.1). These DEGs were mapped to pathways and hallmark gene sets associated with therapy response, providing critical insights into the molecular drivers of systemic and localized immune responses.

To investigate in an unbiased way key molecular pathways associated with immune signaling in blood and TME, we explored chord diagrams generated using iPathway Guide representing gene-pathway interactions (**Fig. 3A and B**). These diagrams highlight connections between pathways, hallmark genes and individual genes, providing insights into shared and unique biological mechanisms in tumor.

**Figure 3.**
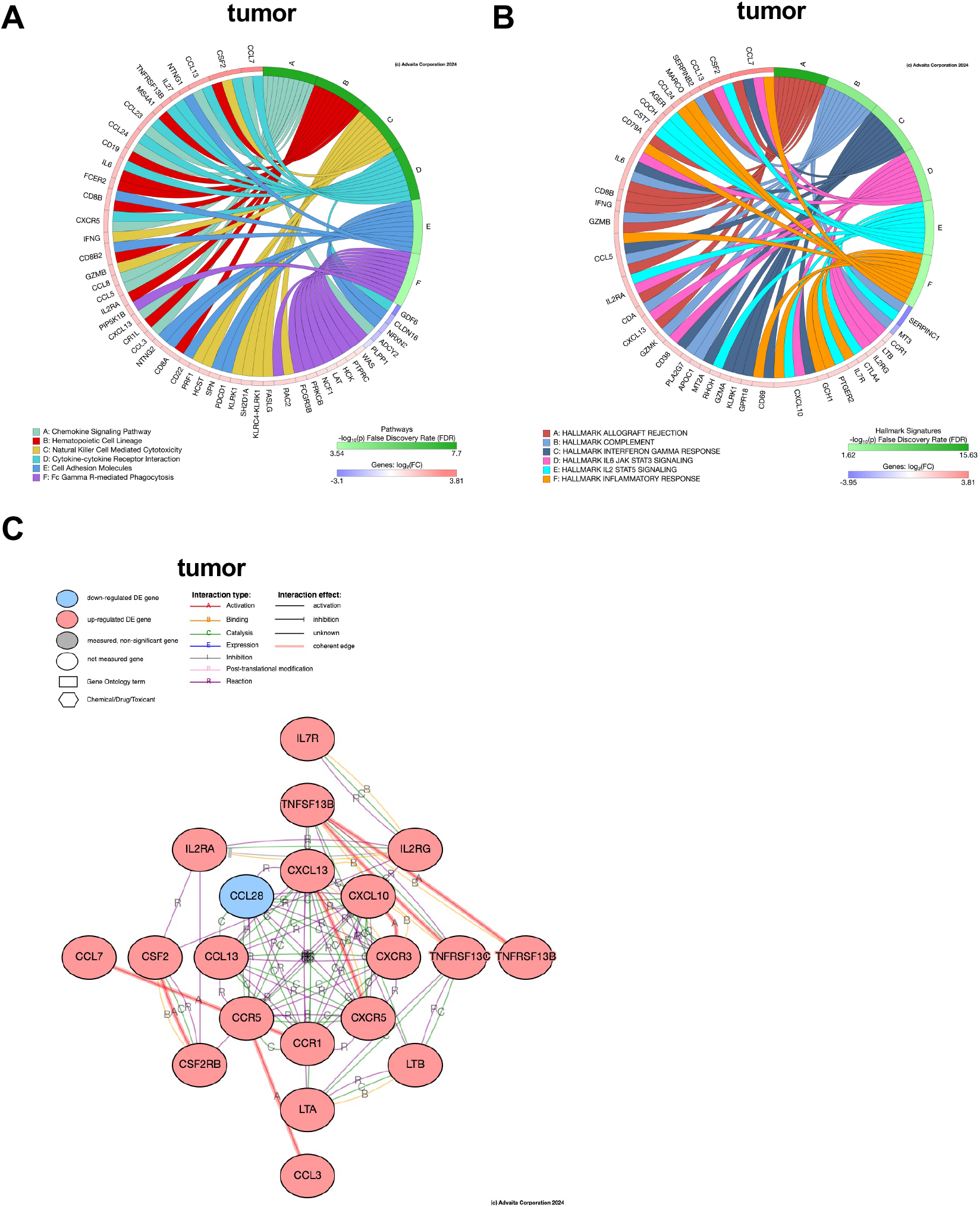
Comprehensive immune pathway analysis in blood and tumor environments. **(A)** Pathway-gene interaction chord diagrams for the TME. The TME network highlights chemokine signaling, Fc gamma receptor-mediated phagocytosis, and cell adhesion molecules, emphasizing immune recruitment and local cytotoxic activity. Nodes are color-coded: red for up-regulated genes and blue for down-regulated genes. **(B)** Heatmaps of hallmark pathway enrichment in the TME. Hallmark pathways like interferon-gamma response, complement activation, and IL6-JAK-STAT3 signaling highlight tumor-specific immune responses. **(C)** Hub gene networks tumor. Hub genes in the TME are characterized by great complexity, features key genes like CXCL13, IL7R, and IL2RA, with predominant up-regulation (red nodes). Interaction types are indicated: activation (green), binding (purple), and reaction (orange). Statistical significance was assessed using gene set enrichment analysis (GSEA) with false discovery rate (FDR) corrections. Thresholds for DEGs ≥1.2-fold change (p < 0.1). This analysis reveals the contrasting complexity and dynamics of immune signaling between localized (tumor) environments.

We next used MSIGDB hallmark signature analysis to investigate in a more stringent way the molecular mechanisms underlying immune regulation in the TME^30^. To this aim, we analyzed hallmark gene signatures using chord diagrams, as shown in **Fig. 3B, left**. Each map illustrates the relationship between hallmark signatures and their associated genes, revealing both shared and unique features of these biological contexts. The hallmark signatures in the TME (**Fig. 3B, right**) include *allograft rejection, complement, interferon gamma response, IL6-JAK-STAT3 signaling, IL2-STAT5 signaling*, and *inflammatory response*. These pathways emphasize the immune-specific processes relevant to tumor immunity. Genes such as *IFNG, CD8A*, and *GZMB* are highly interconnected, reflecting the activation of cytotoxic and inflammatory responses within the tumor microenvironment. Unique to this environment are pathways like complement activation and interferon gamma response, which suggest robust immune signaling, likely triggered by tumor-associated antigens.

We next focused on the common DEGs from TME to generate a thorough interactome with the aim of identifying hub genes that can serve as potential therapeutic targets (**Fig. 3C**). The tumor network was more extensive, consisting of 22 nodes and displaying significantly higher complexity (**Fig. 3C**). This network contained predominantly up-regulated DE genes (red nodes), with *CCL28* as the only down-regulated DE gene (blue node). Central players in the tumor network included *IL7R, IL2RA, IL2RG, TNFSF13B, CXCL13, CXCL10*, and *CCR5*, which form highly interconnected hubs. The network was enriched with activation (A), binding (B), and reaction (R) interactions, particularly involving chemokine and cytokine families such as the *CCL/CXCL* and *IL2/IL7* signaling pathways. The intricate structure of the tumor network suggests a microenvironment reliant on chemokine-mediated immune regulation, likely driving immune cell trafficking and inflammatory responses within the tumor.

The tumor network was enriched for chemokine-driven signaling, reflecting localized immune cell recruitment and activation, while the blood network was more enriched in cytokine-mediated signaling, indicative of broader systemic immune responses. Shared pathways, including *IL2/STAT-5* signaling and inflammatory response suggest crosstalk between systemic and tumor immunity, with cytokines like *IL2* and *IL6* potentially amplifying tumor immune activation and tumor-released chemokines like *CCL5* and *CXCL13* influencing systemic immune recruitment. Distinct pathways, such as *mTORC1* signaling in the blood and complement activation in tumors, highlight environment-specific immune processes.

Furthermore, the tumor network was larger and more densely interconnected than the blood network, reflecting the highly active local immune TME.

This comprehensive and detailed methodology not only provides a unique depth of insight into systemic and localized immune dynamics but also establishes a robust framework for uncovering therapeutic targets and biomarkers of response in mRCC.

## Discussion

In this study, we characterized immune responses in patients with mRCC treated with cICI, specifically ipilimumab (anti-CTLA-4) and nivolumab (anti-PD1). By analyzing both peripheral blood and the TME, we identified systemic immune changes associated with treatment response, notably the enrichment of Th17 CD4+ T cells and their pathways in responders. These findings were corroborated by observations in tumor tissues, underscoring the novel and critical role of Th17 cells in mediating antitumor immunity in mRCC following cICI. This suggests a promising avenue for leveraging Th17 cells as biomarkers and therapeutic mediators in mRCC.

CD4 T cells play a central role in coordinating immune responses, and their functional phenotypes significantly influence the efficacy of immunotherapies in RCC. Recent studies underscore the dynamic interplay between CD4 T cell subsets and the TME, revealing both therapeutic opportunities and challenges. For instance, the shift towards a Th2- and Th17-dominant profile in RCC, coupled with reduced Th1 activity and regulatory T cell (Treg) dysregulation, highlights the immune imbalance in this disease^31^. These changes correlate with tumor progression and underscore the importance of re-establishing a functional Th1/Th2 balance for effective antitumor responses.

The emerging role of Th17 cells in driving antitumor immunity, particularly in the context of ICI, has been a focal point of recent investigations. Th17 CD4+ T cells play a complex role in cancer, as they are capable of both promoting and inhibiting tumor progression depending on the context and tumor type^32-34^. For instance, Th17 cells and IL-17 have been implicated in early oncogenesis by inducing cancer stem cell renewal^35^ and supporting myeloid-derived suppressor cells via IL-6/IL-23/IL-1b^36^, which can promote tumor growth. Conversely, Th17 cells have been associated with antitumor immunity in more advanced tumors^34^. Circulating Th17 cells correlate with better prognosis in chronic lymphocytic leukemia patients^37^ and have been linked to improved responses to ICI therapy^19,38^. Preclinical studies also support this role: Th17 cells inhibited tumor growth in mouse models of lymphoma, mastocytoma, by activating CD8+ T cells and dendritic cells^39-41^. Importantly, Th17-related inflammation must be carefully regulated as excessive activity can also support tumor progression under specific conditions. Conversely, targeting suppressive pathways, such as those mediated by CCR4, has demonstrated potential to remodel the TME in RCC by increasing Th1-associated cytokines and NK cell infiltration, further emphasizing the reliance of these effects on adaptive immunity and CD4 T cells^42^.

Additionally, novel immune signatures involving CD4 T cells, such as those identified through m5C RNA methylation-related pathways or Th17-centric transcriptional profiles, have been linked to better therapeutic outcomes and could guide patient stratification and therapy optimization^43^. The balance of naïve and effector CD4 T cell subsets, including Th1, Th2, and Th17 populations, also appears to play a critical role in response to ICI, as observed in clinical trials and immune profiling studies^44^.

Future research should build on our findings by further exploring the critical role of CD4 T cell subsets, particularly Th17 cells, in mediating antitumor responses in RCC. Our study demonstrated a Th17-centric signature in both peripheral blood and the TME of responders to cICI, highlighting the importance of Th17 cells in driving effective therapeutic outcomes. This suggests the potential of targeting Th17-associated pathways, such as IL-21 signaling, to enhance immune responses in non-responders. Additionally, the observed delayed transition of CD4 T cells from naïve to Th17 phenotypes in non-responders underscores the need to investigate mechanisms of immune dysregulation and identify strategies to accelerate or restore this transition.

Future studies should also focus on integrating minimally invasive approaches, such as liquid biopsies, with tumor transcriptomic analyses to track dynamic changes in CD4 T cell phenotypes over the course of treatment. This could provide real-time insights into therapeutic efficacy and resistance mechanisms. Moreover, combination strategies targeting suppressive pathways, such as CCR4, alongside cICI, may enhance the immune response by reshaping the TME to favor Th1 and Th17-driven immunity. These efforts should also evaluate the therapeutic potential of targeting key genes and pathways identified in our study, including IL13, IFNG, and IL2, which were enriched in responder Th17 cells.

Ultimately, leveraging these findings could lead to the development of novel biomarkers and combination therapies that improve patient stratification, overcome resistance, and maximize the efficacy of immunotherapy in RCC.

A notable strength of our work lies in the analysis of systemic and tumor specific immune responses using a systematic multi-omic approach combined with multiple analysis pipelines that enabled a comprehensive study of the immunologic changes that occur following treatment and that correlate with response to cICI.

Studies on the immune landscape of the TME in mRCC have largely focused on PD-L1 expression and effector T cell gene signatures, which have underperformed as stratification tools^5,14^.

Similarly, tumor mutational burden (TMB) is low in RCC and fails to predict clinical outcomes^5,14^ and CD8+ T cell infiltration, despite notable, does not consistently correlate with improved outcomes in mRCC^29^.

More recent unbiased approaches have shown that high T cell/low myeloid infiltration and high B cell abundance are enriched in responders to ICI therapy^16,17^. For instance, multiomic analyses have also reported factors that are associated with response to ICI in patients with mRCC^45,46^. However, comprehensive and systematic definitions of immune correlates remain elusive, partly due to the cost, time, and clinical complications associated with serial tumor biopsies.

As a result, no clear biomarker has been established so far that is commonly used in clinics to differentiate between patients that respond and patients that don’t respond to ICI treatment^47^. The analysis of tissues from the IMmotion150, a phase 2 study comparing sunitinib to atezolizumab and bevacizumab treatment, showed that signatures associated with angiogenesis correlated with improved outcome in patients treated with sunitinib^17^. Furthermore, T cell effector and IFNγ signatures were linked to better progression-free survival in patients receiving ICI^17^. In another study, there was also an immune signature associated with response in patients with metastatic ccRCC receiving ICI^45^. Interestingly, while not specified in detail, durable responses and prolonged survival suggest an association with specific immune signatures activated by dual checkpoint inhibition therapy in the Checkmate214 study. Similar to the B cell changes we have observed, a B cell signature was linked to the response to ipilimumab and nivolumab. In recent work by Kinget and colleagues, patients with a repertoire of HLAs that were more restricted to neoantigens had a better response to ICI therapy^46^.

The identification of Th17-centric pathways in responders that we report in the present study has significant translational implications. Th17 cells, IL-21, and associated pathways could serve as predictive biomarkers for stratifying patients likely to benefit from ICI therapy. Additionally, these pathways may present therapeutic targets to enhance antitumor immunity, particularly in non-responders. The observed upregulation of dendritic cells and B cells in responders further supports the potential for combinatorial approaches to amplify immune responses^45,48^. Furthermore, our results strongly support the concept that minimally invasive liquid biopsies, in combination with tumor biopsies, could bridge existing gaps in biomarker discovery and improve response stratification in mRCC.

### Limitations, Strengths, and Future Directions

While our study involved a relatively small cohort, this challenge is shared across the field of mRCC research due to the rarity and complexity of sequential tumor and blood sampling. However, the comprehensive and detailed nature of our analysis stands out, particularly in the use of serial blood and primary and metastatic tumor samples from the same patients. This approach aligns with the **reverse translational framework**, where patient samples are utilized to elucidate immune responses, which are then tested in preclinical models. This method provides critical insights into how tumor-immune interactions evolve and can guide the rational design of future therapeutic strategies^49^.

The high purity of the cell populations analyzed in our study further enhances the robustness and specificity of our findings. Similar to the systematic immune monitoring discussed by Sharma and Allison, our study aims to leverage detailed immune profiling to unravel the mechanisms of resistance to treatment and identify new therapeutic targets^49^. Through integrating high-dimensional technologies like single-cell RNA sequencing and multiplexed imaging, we could push the boundaries of our current understanding, as seen in recent advances that map the tumor microenvironment and immune responses in unprecedented detail.

Future studies should build on these insights by validating our results in larger, independent cohorts, and further exploring the mechanistic roles of **Th17 cells** across other cancer types, as well as the evolving immune landscapes. Given that therapy with ICI has shown varying efficacy across different cancers, understanding immune profiles—especially **myeloid subsets** and **tertiary lymphoid structures**—can be pivotal in predicting patient outcomes.

Additionally, integrating **minimally invasive liquid biopsies** with tumor transcriptomics holds great promise for advancing comprehensive immune monitoring and improving response stratification in mRCC. As demonstrated in the reverse translational approach, combining tumor tissue data with longitudinal immune profiling can provide actionable insights for developing targeted therapies that overcome resistance mechanisms. Future studies that adopt this integrated approach could potentially transform the landscape of mRCC treatment, leading to better patient stratification and improved clinical outcomes.

### Conclusions

Our study provides a significant step forward in understanding immune correlates of response to dual ICI therapy in mRCC. By identifying Th17-centric pathways and bridging systemic and tumor-specific immune dynamics, we offer a robust framework for the development of predictive biomarkers and therapeutic strategies to enhance immunotherapy outcomes. These findings underscore the potential for advancing personalized treatment approaches in mRCC and beyond.

## Data Availability

All data produced in the present study are available upon reasonable request to the authors

## Funding

This work was supported and funded by Bristol-Myers Squibb, the Krebsliga Schweiz, and from the Schoenemakers-Müller Foundation to H.L., ACS-IRG and DoD HT94252510707 to C.K., American Cancer Society RSG-24-1255025-01-IBCD and NCI R01CA258882 to S.G. The work was supported in part by the NCI through the Cancer Bioinformatics Core, UPMC Hillman Cancer Center, University of Pittsburg (P30CA047904), and the Cell Evaluation & Therapy Shared Resource, Hollings Cancer Center, Medical University of South Carolina (P30 CA138313).

## Conflict of interest

H.L. received travel grants and consultant fees from Bristol-Myers Squibb (BMS) and Merck, Sharp and Dohme (MSD). H.L. received research support from BMS, Novartis, GlycoEra and Palleon Pharmaceuticals. F.S. received travel grants from Roche, BMS, Celgene and research support from BMS and Takeda. C.K. declares ownership of Cell X Analytics LLC and contributions of reagents from MSD.

## Authors Contributions

Following the established authorship criteria (Committee on Publication Ethics - COPE), the contributions of each author to the study are as follows: A.B. collected study material, data and supported the clinical study. G.M. analyzed bulk RNA seq data and wrote the manuscript. P.H. performed critical data analysis. A.Z. contributed to the conception and design of the study. L.B. served as the primary pathologist, contributing to patient material collection, the acquisition and interpretation of. R.I. performed RNA-seq analysis, contributing to data acquisition and interpretation. H.L. and F.S. collaboratively designed and planned the clinical trial, were integral to the conception and design of the study, patient treatment, supported by the SAKK (Swiss Academy of Clinical Research) and sponsored by Bristol-Myers Squibb. H.L. and C.K. supervised the study. S.G., C.K., and H.L. drafted and revised the manuscript. S.G. and C.K. performed CyTOF contributing to data acquisition. S.G., A.S., S.O., and C.K. analyzed CyTOF data, while A.S., S.O., and M.D.R. designed and executed the data analysis pipeline and generated figures for CyTOF. T.C. and G.H. performed system biology analysis of the RNA sequencing data. All authors approved the submitted version of the manuscript and any subsequent substantially modified versions.

## Acknowledgement

The authors extend their deepest gratitude to the patients, families, and veterans whose courage and commitment made this study possible. We believe that by joining our passion as researchers with your dedication as patients and advocates, we can move toward a better future in kidney cancer treatment. “No one is an island” and in this shared journey, we also acknowledge the vital role of advocacy organizations such as KidneyCAN.org, whose tireless efforts ensure that the voices of patients and families guide research and progress.

## Code and data availability

The code to reproduce the Cytof analysis is freely available on Github: Raw data for CyTOF are available

## Notes

### Clinical Trial

NCT03297593

### Funding Statement

This work was supported and funded by Bristol-Myers Squibb, the Krebsliga Schweiz, and from the Schoenemakers-Mueller Foundation to H.L., ACS-IRG and DoD HT94252510707 to C.K., American Cancer Society RSG-24-1255025-01-IBCD and NCI R01 CA258882 to S.G. The work was supported in part by the NCI through the Cancer Bioinformatics Core, UPMC Hillman Cancer Center, University of Pittsburg (P30CA047904), and the Cell Evaluation & Therapy Shared Resource, Hollings Cancer Center, Medical University of South Carolina (P30 CA138313).

### Author Declarations

The Institutional Review Board of the Medical University of South Carolina waived ethical approval for this work, as the study was determined to meet the criteria for Not Human Research (NHR; Pro00144924). This study involves the analysis of pre-existing, biobanked clinical samples obtained from a completed Phase II clinical trial (NCT03297593) conducted in Switzerland under the oversight of the Ethics Committee of Northwestern and Central Switzerland (EKNZ).

