## Supplement for "A Th17 signature correlates with response to dual CTLA-4 and PD-1 blockade in metastatic renal cell carcinoma"

### Supplementary Data

*Sample preparation.*

PBMC were isolated via Ficoll (GE Healthcare) density gradient centrifugation from fresh blood and cryopreserved in fetal bovine serum (VWR) plus 10% DMSO (Sigma). All cells were finally stored in liquid nitrogen. On the day of the experiment, PBMC were thawed dropwise added to RPMI containing 5% FCS and 100U of DNase (Sigma), incubated at room temperature for 10min, centrifuged, filtered through a 40microm mesh, and counted. Each sample was enriched using custom anti-human TCR-alpha (clone: IP26, BioLegend) magnetic bead selection (StemCell Technologies) according to the manufacturer's instructions into one T cell and one fraction of non-T-cells.

*Heavy metal conjugation of antibodies.*

Conjugation of anti-human antibodies to heavy metal isotopes of the lanthanide series was conducted using the MaxPar X8 antibody-labeling kit (Standard BioTools) following the manufacturer's recommendations. Where available, pre-conjugated antibodies were obtained from Standard BioTools (**Supp. Tables 2 and 3**). Protein content was assessed by NanoDrop (ThermoFisher) and an antibody stabilizer (Candor) was added to a final volume of 50% and the conjugated antibody was stored at 4°C.

*Bulk RNA sequencing analysis*

Reads were aligned to the human genome with default parameters. Read and alignment quality was evaluated using the qQCReport function of the Bioconductor package QuasR (v 1.30.0). The featureCounts function from Bioconductor package Rsubread (v 2.4.3) was used to count the number of read (5'ends) overlapping with the exons of each gene assuming an exon union model (with used gene model provided by ensembl v101). The data were normalized by applying the TMM method from Bioconductor edgeR package (version 3.32.1). Only genes having log2 CPM counts bigger than 0 in at least 2 samples were kept for the further analyses. The principal component analysis was based on 25% of most variable genes in the dataset. The differentially expressed genes were identified using the quasi-likelihood (QL) method implemented in edgeR package (version 3.32.1) using replicate id as covariate. Genes with FDR smaller than 0.05 and either positive or negative log2 fold change were considered as differentially expressed. The DEGs found with the CITE-Seq analysis from the CD4 EM, CD4 EF, CD4 Th17, Monocytes, B cells, and NK cells were used for generating an enrichment score. The GEP_MSD2018 signature (CCL5, CD27, CD274, CD276, CD8A, CMKLR1, CXCL9, CXCR6, HLA-DQA1, HLA-DRB1, HLA-E, IDO1, LAG3, NKG7, PDCD1LG2, PSMB10, STAT1, TIGIT) was also included for comparison. The enrichment score was calculated with the Gene Set Variation Analysis (GSVA) package with default parameters on log2-transformed Transcript Per Million (TPM) values. Heatmaps were generated with the pheatmap package.

*Statistics*

Patient characteristics are presented as descriptive analysis. For statistical considerations, GraphPad Prism was used (Version 8). Differential abundance analysis of CyTOF data was performed with the voom-diffcyt method[22], which uses the limma package[50] to calculate moderated tests at the cluster level and the voom method to normalize cluster cell counts[51]. For CITE-Seq data, the differential state analysis was performed using the pseudo-bulk method implemented by the muscat package[52], using the generalized linear model implemented by edgeR[53] for the statistical testing. The differential abundance was performed using the same approach, performed on the number of cell counts per label, also implemented by edgeR.

### Supplementary Figures.

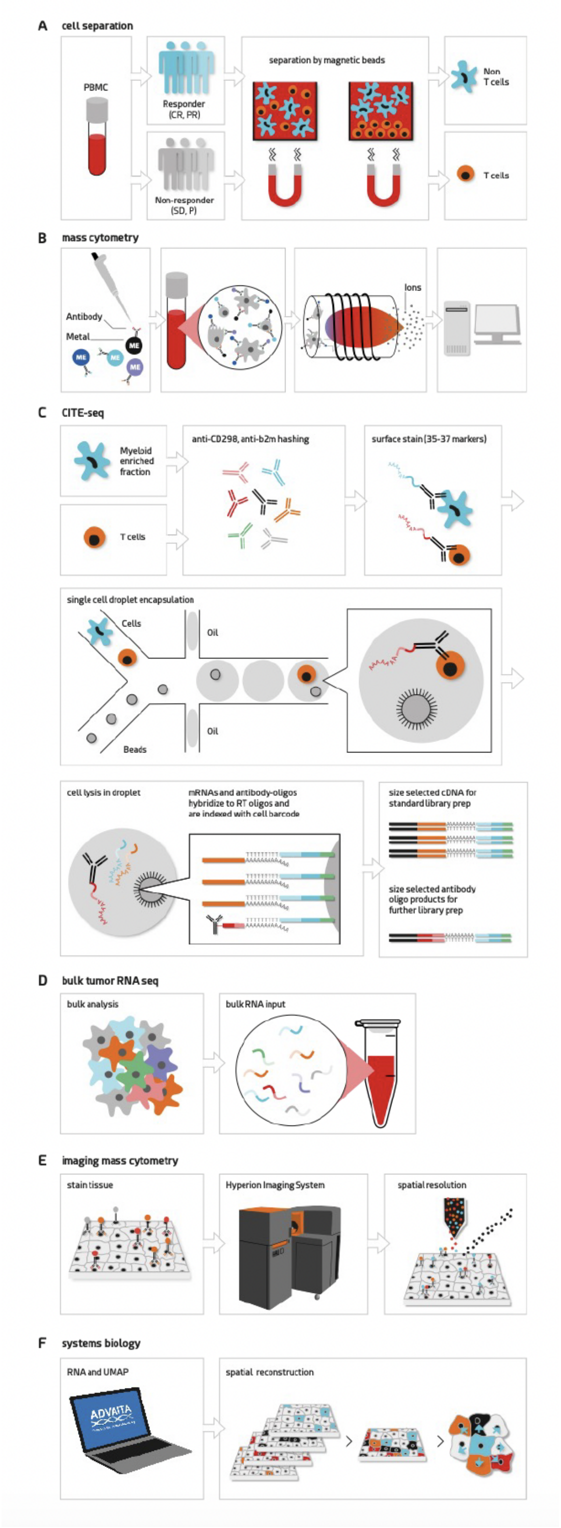


**Supplementary Figure 1. Graphical Summary of study workflow. A**, Schematic workflow illustration of the analysis of PBMC collected from the CA209-980 trial, SAKK07/17 trial. Patients were grouped after iRECIST criteria into responders (n= 12; CR=complete response, PR= partial response) and non-responders (n=3; SD=stable disease, P=progressive disease). Enrichment of T cell and myeloid cell populations from patients using magnetic beads. **B**, Further processing of enriched cell populations using CITE-Seq. Enriched cell solutions are barcoded using hashing, pooled, and surface stained with DNA tagged antibodies. Cell solutions are then subjected to single cells sequencing. Individual cells are linked to bound surface antibodies and single-cell mRNA by unique beads. Subsequently, cell types are identified from antibody oligo libraries and related to correlating gene expression profiles. Samples were analyzed prior to treatment initiation and at the time of RECIST evaluation, typically conducted 12 weeks after the commencement of combination immune checkpoint inhibitor (cICI) therapy.

**A**

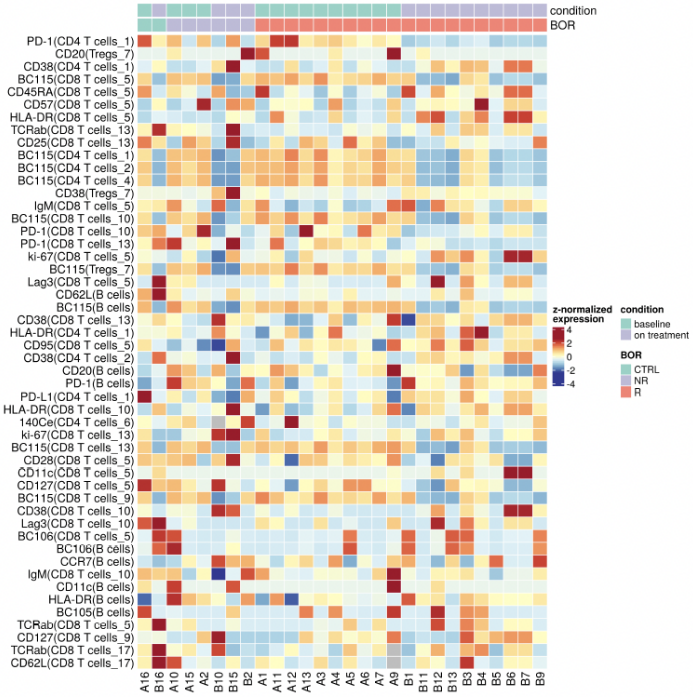


**B**

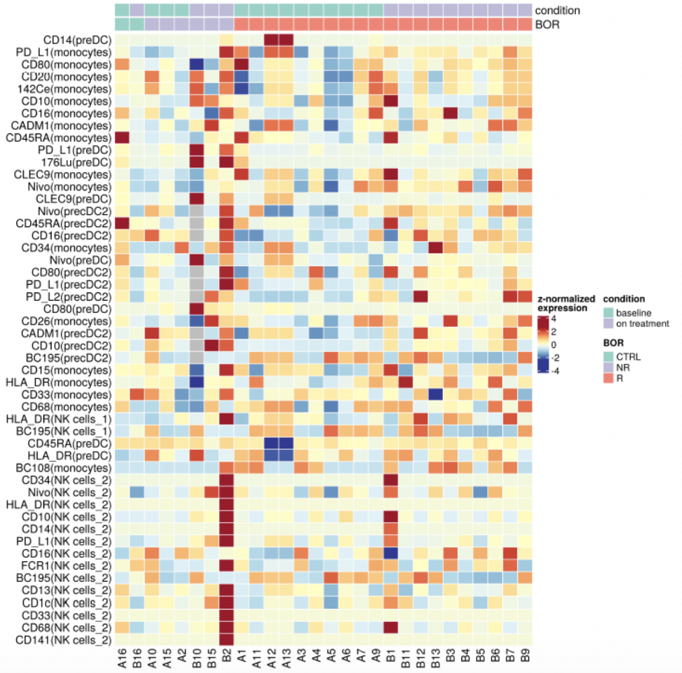


**Supplementary Figure 2 Differences in protein surface expression between groups by mass cytometry. A**, T cell fraction. **B**, non-T-cell fraction.

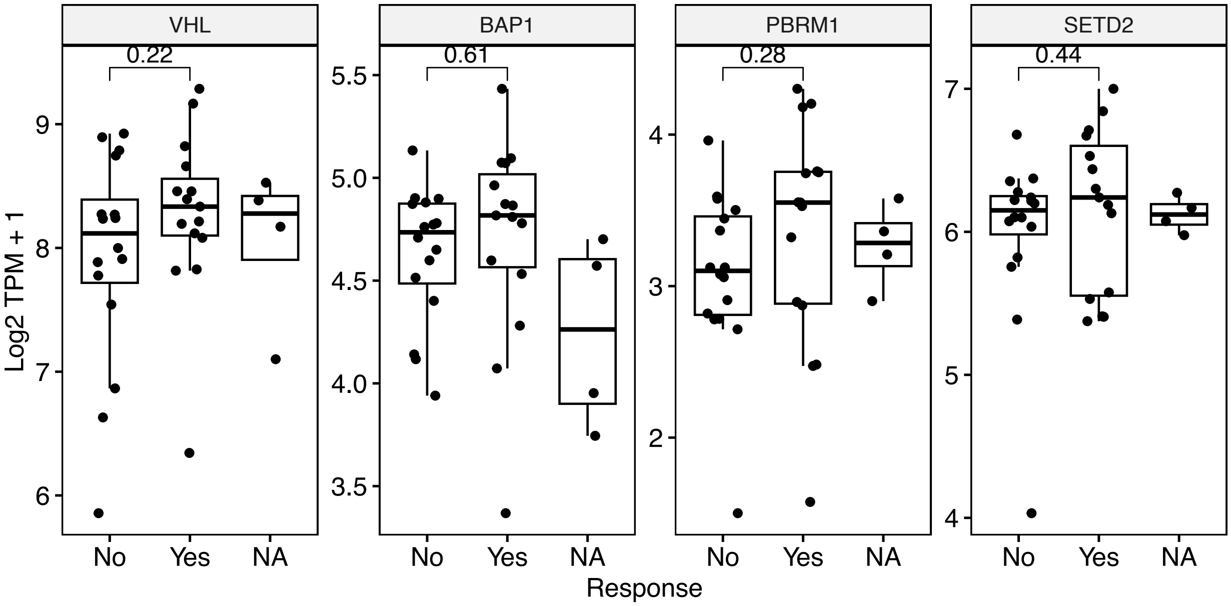


**Supplemental Figure 3. Gene expression of VHL, BAP1, PBRM1, and SETD2 in relation to response to immune checkpoint inhibition in clear cell renal cell carcinoma.** Gene expression data for four genes (VHL, BAP1, PBRM1, SETD2) in relation to response to immune checkpoint inhibition (ICI) in clear cell renal cell carcinoma (ccRCC). Gene expression is shown as log2 TPM + 1, stratified by ICI responsiveness ("Yes" for response, "No" for no response, "NA" for unavailable data). The x-axis represents response status, while the y-axis shows gene expression levels. Numbers on brackets refer to p-values.

| Mass | Metal | Marker | Clone | RRID | Cat# | Vendor |
|---|---|---|---|---|---|---|
| **Technical** | | | | | | |
| 105 | Pd | CD298+b2m | LNH-94; 2M2 | AB_2876646; AB_492835 | 341712, 316302 | BioLegend |
| 106 | Pd | CD298+b2m | LNH-94; 2M2 | AB_2876646; AB_492835 | 341712, 316302 | BioLegend |
| 108 | Pd | CD298+b2m | LNH-94; 2M2 | AB_2876646; AB_492835 | 341712, 316302 | BioLegend |
| 113 | In | CD298+b2m | LNH-94; 2M2 | AB_2876646; AB_492835 | 341712, 316302 | BioLegend |
| 115 | In | CD298+b2m | LNH-94; 2M2 | AB_2876646; AB_492835 | 341712, 316302 | BioLegend |
| 194 | Pt | CD298+b2m | LNH-94; 2M2 | AB_2876646; AB_492835 | 341712, 316302 | BioLegend |
| 195 | Pt | CD298+b2m | LNH-94; 2M2 | AB_2876646; AB_492835 | 341712, 316302 | BioLegend |
| 196 | Pt | CD298+b2m | LNH-94; 2M2 | AB_2876646; AB_492835 | 341712, 316302 | BioLegend |
| 198 | Pt | CD298+b2m | LNH-94; 2M2 | AB_2876646; AB_492835 | 341712, 316302 | BioLegend |
| 191 | Ir | DNA1 | NA | NA | 201192B | Standard Biotools |
| 193 | Ir | DNA2 | NA | NA | 201192B | Standard Biotools |
| 103 | Rh | Dead | NA | NA | 201103B | Standard Biotools |
| **Lineage markers** | | | | | | |
| 89 | Y | CD45 | HI30 | AB_2938863 | 3089003B | Standard Biotools |
| 146 | Nd | CD8 | RPA-T8 | AB_2687641 | 3146001 | Standard Biotools |
| 151 | Eu | CD14 | M5E2 | AB_2810244 | 3151009B | Standard Biotools |
| 152 | Sm | TCRgd | 11F2 | AB_2687643 | 3152008B | Standard Biotools |
| 163 | Dy | CD33 | WM53 | AB_2687857 | 3163023 | Standard Biotools |
| 171 | Yb | CD20 | 2H7 | AB_2802112 | 3171012B | Standard Biotools |
| 170 | Er | TCRab | IP26 | AB_2738921 | 564728 | BD |
| 164 | Dy | CD95 | DX2 | AB_2858235 | 3164008B | Standard Biotools |
| 145 | Nd | CD4 | RPA-T4 | AB_2687832 | 3145002B | Standard Biotools |
| 144 | Nd | IgM | MHM-88 | AB_493003 | 314502 | BioLegend |
| 149 | Sm | CD66b | 80H3 | AB_960677 | NB100-64916 | Novus |
| 155 | Gd | CD56 | B159 | AB_2861412 | 3155008B | Standard Biotools |
| 166 | Er | NKP46 | 9E2 | AB_2149297 | 557847 | BD |
| **Migration and differentiation** | | | | | | |
| 141 | Pr | CCR6 | G034E3 | AB_2687639 | 3141003A | Standard Biotools |
| 142 | Nd | CD11a | HI111 | AB_2877095 | 3142006B | Standard Biotools |
| 143 | Nd | CD45RA | HI100 | AB_2651156 | 3143006B | Standard Biotools |
| 147 | Sm | CD11c | Bu15 | AB_2687850 | 3147008B | Standard Biotools |
| 153 | Eu | CD62L | DREG56 | AB_2810245 | 3153004B | Standard Biotools |
| 156 | Gd | CXCR3 | G025H7 | AB_2687646 | 3156004B | Standard Biotools |
| 158 | Gd | CCR4 | 205410 | AB_2893003 | 3158032A | Standard Biotools |
| 159 | Tb | CCR7 | G043H7 | AB_2714155 | 3159003 | Standard Biotools |
| 160 | Gd | CD28 | CD28.2 | AB_2868400 | 3160003B | Standard Biotools |
| 162 | Dy | FoxP3 | PCH101 | AB_2938865 | 3162024A | Standard Biotools |
| 165 | Ho | CD45RO | UCHL1 | AB_275642 | 3165011B | Standard Biotools |
| 171 | Yb | Grz_B | GB11 | AB_2687652 | 3171002B | Standard Biotools |
| 172 | Yb | CD57 | HCD57 | AB_2888930 | 3172009B | Standard Biotools |
| 173 | Yb | HLA-DR | L243 | AB_2810248 | 3173005B | Standard Biotools |
| 174 | Yb | CD94 | HP-3D9 | AB_2756429 | 3174015B | Standard Biotools |
| 176 | Yb | CD127 | A019D5 | AB_2687863 | 3176004 | Standard Biotools |
| 209 | Bi | CD11b | ICRF44 | AB_2687654 | 3209003 | Standard Biotools |
| **Immune modulation** | | | | | | |
| 167 | Er | CD38 | HIT2 | AB_2802110 | 3167001B | Standard Biotools |
| 168 | Er | ki-67 | Ki67 | AB_2810856 | 3168001B | Standard Biotools |
| 175 | Lu | PD_L1 | 29E.2A3 | AB_2687638 | 3175017B | Standard Biotools |
| 148 | Nd | Lag-3 | 11C3C56 | AB_2616876 | 369302 | BioLegend |
| 154 | Sm | Tim-3 | F38-2E2 | AB_2893002 | 3154010B | Standard Biotools |
| 161 | Dy | CTLA4 | 14D3 | AB_2687649 | 3161004B | Standard Biotools |
| 169 | Tm | CD25 | 2A3 | AB_2938861 | 3169003B | Standard Biotools |
| 174 | Yb | PD-1 | EH12.2H7 | AB_286840 | 3174020B | Standard Biotools |

**Supplementary Table 1 - CyTOF T cell panel.**

| Mass | Metal | Marker | Clone | RRID | Cat# | Vendor |
|---|---|---|---|---|---|---|
| **Technical** | | | | | | |
| 105 | Pd | CD298+b2m | LNH-94; 2M2 | AB_2876646; AB_492835 | 341712, 316302 | BioLegend |
| 106 | Pd | CD298+b2m | LNH-94; 2M2 | AB_2876646; AB_492835 | 341712, 316302 | BioLegend |
| 108 | Pd | CD298+b2m | LNH-94; 2M2 | AB_2876646; AB_492835 | 341712, 316302 | BioLegend |
| 113 | In | CD298+b2m | LNH-94; 2M2 | AB_2876646; AB_492835 | 341712, 316302 | BioLegend |
| 115 | In | CD298+b2m | LNH-94; 2M2 | AB_2876646; AB_492835 | 341712, 316302 | BioLegend |
| 194 | Pt | CD298+b2m | LNH-94; 2M2 | AB_2876646; AB_492835 | 341712, 316302 | BioLegend |
| 195 | Pt | CD298+b2m | LNH-94; 2M2 | AB_2876646; AB_492835 | 341712, 316302 | BioLegend |
| 196 | Pt | CD298+b2m | LNH-94; 2M2 | AB_2876646; AB_492835 | 341712, 316302 | BioLegend |
| 198 | Pt | CD298+b2m | LNH-94; 2M2 | AB_2876646; AB_492835 | 341712, 316302 | BioLegend |
| 191 | Ir | DNA1 | NA | NA | 201192B | Standard Biotools |
| 193 | Ir | DNA2 | NA | NA | 201192B | Standard Biotools |
| 103 | Rh | Dead | NA | NA | 201103B | Standard Biotools |
| **Lineage Markers** | | | | | | |
| 141 | Pr | IgM | MHM-88 | AB_493003 | 314502 | BioLegend |
| 142 | Nd | CD20 | 2H7 | AB_46715 | 302302 | BioLegend |
| 145 | Nd | CD7 | CD7-6B7 | AB_3106934 | 3145013B | Standard Biotools |
| 148 | Nd | CD16 | 3G8 | AB_2661791 | 3148004B | Standard Biotools |
| 149 | Sm | CD66b | 80H3 | AB_960677 | NB100-64916 | NOVUS |
| 153 | Eu | CD192 (CCR2) | K036C2 | AB_3106935 | 3153023B | Standard Biotools |
| 156 | Gd | CD183 (CXCR3) | GO25H7 | AB_2687646 | 3156004B | Standard Biotools |
| 158 | Gd | CD10 | HI10a | AB_2921314 | 3158011B | Standard Biotools |
| 160 | Gd | CD14 | M5E2 | AB_2661801 | 3160001B | Standard Biotools |
| 166 | Er | CD34 | 581 | AB_2756424 | 3166012B | Standard Biotools |
| 169 | Tm | CD33 | WM53 | AB_2802111 | 3169010B | Standard Biotools |
| 170 | Er | HLA-DR | L243 | AB_2888929 | 3170013B | Standard Biotools |
| 171 | Yb | CD68 | Y1/82A | AB_2687637 | 3171011B | Standard Biotools |
| 172 | Yb | CX3CR1 | 2A9-1 | | 17-6039-42 | ThermoFisher |
| 176 | Yb | CD56 | NCAM16.2 | AB_2661813 | 3176008B | Standard Biotools |
| 209 | Bi | CD11b | ICRF44 | AB_2687654 | 3209003B | Standard Biotools |
| **DC Differentiation** | | | | | | |
| 173 | Yb | CD141 (BDCA3) | 1A4 | AB_2714156 | 3173002B | Standard Biotools |
| 152 | Sm | CADM1 | 3E1 | AB_592783 | CM004-3 | MBL |
| 161 | Dy | CLEC9A | 8F9 | AB_2810252 | 3161018B | Standard Biotools |
| 164 | Dy | CD26 | BA5b | AB_314286 | 302702 | BioLegend |
| 154 | Sm | CD1c | L161 | AB_1088995 | 331502 | BioLegend |
| 146 | Nd | CD11c | 3.9 | AB_3106934 | 3146014B | Standard Biotools |
| 150 | Nd | CD64 (FCR1) | AER-37 (CRA-1) | AB_3106937 | 3150027B | Standard Biotools |
| 151 | Eu | CD2 | TS1/8 | AB_3106938 | 3151003B | Standard Biotools |
| 143 | Nd | CD123 | 6H6 | AB_2811081 | 3143014B | Standard Biotools |
| 155 | Gd | CD45RA | HI100 | AB_2810246 | 3155011b | Standard Biotools |
| 159 | Tb | CD303 (BDCA2/CLEC4C) | 201A | AB_2563739 | 354202 | BioLegend |
| **Immune Activation** | | | | | | |
| 144 | Nd | CD15 | W6D3 | AB_2892685 | 3144019B | Standard Biotools |
| 147 | Sm | CD13 | WM15 | AB_3106939 | 3147014B | Standard Biotools |
| 162 | Dy | CD86 | 2D10.4 | AB_2687856) | 3162010B | Standard Biotools |
| 163 | Dy | CD272 (BTLA) | MIH26 | AB_2910546 | 3163009B | Standard Biotools |
| 167 | Er | CD38 | HIT2 | AB_2802110 | 3167001B | Standard Biotools |
| 168 | Er | CD274 (anti-PD-1, Nivolumab) | 5C4.B8 | AB_2893893 | NA | BMS |
| 172 | Yb | CD273 (PD-L2) | 1054620 | AB_3657532 | MAB11351 | RnD Systems |
| 175 | Lu | CD274 (PD-L1) | 29E.2A3 | AB_2687638 | 3175017B | Standard Biotools |

**Supplementary Table 2 - CyTOF non-T cell panel.**

**Supplementary Table 3 - Clinical characteristics of responders (R) and non-responders (NR)**

| Characteristic | responder (N = 16) | non-responder (N = 8) |
|---|---|---|
| **Median age — yr (range)** | 72 (45–84) | 61 (49–74) |
| **Male sex — no. (%)** | 13 (81.3) | 7 (87.5) |
| **Years from diagnosis (range)** | 2.05 (0-9.1) | 0.7 (0-3.4) |
| **Site of tissue — no. (%)** | | |
| kidney | 4 (25) | 1 (12.5) |
| lung | 6 (37.5) | 2 (25) |
| other | 6 (37.5) | 5 (62.5) |
| **Laboratory values** | | |
| hemoglobin (g/L) | 131 | 119 |
| thrombocytes (G/L) | 306 | 341 |
| granulocytes (G/L) | 5.16 | 5.28 |
| LDH (U/L) | 259.4 | 248.5 |
| CRP | 47.7 | 25.5 |
| Calcium (mMol) | 2.35 | 2.26 |
